# Information Sharing through Digital Service Agreement

**DOI:** 10.1101/2023.01.18.23284718

**Authors:** Luis B Elvas, Berit Helgheim, João CA Ferreira

## Abstract

Data sharing and services reuse in the health sector is a significant problem due to privacy, and security issues. The European Commission has classified health data as a unique resource owing to the ability to do both prospective and retrospective research at a low cost. Similarly, the OECD encourages member nations to create and implement health data governance systems that protect individual privacy while allowing data sharing. This paper aimed to describe a conceptual framework to allow medical information sharing among health entities in a secure environment. A framework of shared Artificial Intelligent services is proposed to provide a safe environment for information sharing based on digital services agreements (DSA) and a shared services infrastructure for artificial intelligence (AI) and knowledge creation: From the collaborative platform with privacy, health data can be shared, and shared analytics services will allow an easy and fast application of AI algorithms. The framework allows data prosumers (producers/consumers) to easily express their preferences on sharing their data, which analytics operations can be performed on such data, and by whom the resulting data can be shared, among other relevant aspects. This entails a framework that combines several technologies for expressing and enforcing data-sharing agreements and technologies to perform data analytics operations compliant. Among these technologies, we can mention data-centric policy enforcement mechanisms and data analysis operations directly performed on encrypted data provided by multiple prosumers. The framework is mainly based on an Information Sharing Infrastructure (ISI) and an Information Analysis Infrastructure (IAI) that can be deployed in several ways and on several devices (from cloud to mobile devices).

## Introduction

Healthcare data integration is a vital investigation topic involving patient privacy issues and dealing with different information systems. There is already some work under this topic [1] and [2]; nevertheless, isolated data from a single source is insufficient. Data must be enriched by adding further information (metadata) and integrating it with other data sources. This is especially true in the clinical domain, where other diseases and diagnostics are essential for the correct diagnosis of an illness and preventing worsening conditions. Indeed, integrating and analysing the vast number of data sources and information gathered has been identified as one of the main challenges that need to be dealt with before personalised medicine can be effectively deployed [3]. Integrating data for the analysis implies adequately correlating readings that provide different perspectives of the same object, enabling more complete and detailed analysis and better understanding for the analyst. This integration is often challenging, consuming considerable time and becoming a bottleneck for real-time data access. Due to privacy and data protection, sharing information is a problem because due trust and interoperability problems among institutions [4]. One approach to deal with trust issues between institutions is the use of blockchain [5], nevertheless, has been demonstrated to be a complex solution to implement and will not address the problems that the health-care industry is facing, in fact, it may cause more problems than it solves. One other approach to deal with trust issues among different entities is the Data Sharing Agreements (DSAs) [6]. A DSA is an agreement between two or more parties that want to communicate data in various domains and contexts: it specifies which data to use, for what goals, and how to use it. Essentially, the purpose of DSA is to record the data-sharing regulations that limit both data producers and data consumers and manage the flow of data between them. DSAs are written documents. These can express privacy preferences or contractual requirements for providing and consuming information (i.e., notification of data leakage). The information can often be analysed globally (in the cloud) or locally (in edge devices). Since information sharing is a major issue in health care, our research approach handles the following key components:

1. Information sharing: share information (including security ones) in a controlled manner, ensuring the respect of regulation and confidentiality and integrity both in rest and in transit;
2. Information analytics: advanced analytics functions and engines for data analytics and correlation identifying threats that hide in the massive usage of services and the related amount of logs
3. Mixture of technologies to enable a confidential and collaborative analysis of data: including homomorphic encryption: making computation in a personal and distributed manner;
4. Advanced seamless access mechanisms that take advantage of the analytics and sharing infrastructure to provide continuous authentication, authorisation, and privacy-aware service as privacy-aware data usage control.

This is aligned with the current mobility of doctors among hospitals (private vs public) and patient mobility. Recent research with hospitals is to integrate health data to extract knowledge and information analytics, enabling security as a service to be easily deployed by communities of prosumers. The service allows federations of prosumers and patients to: 1) Have interoperability & portability: between collaborating prosumers that agree to exchange and share events data; 2) Manage incident notification according to law, regulatory aspects as well as contractual agreements; and 3) Experience improved business intelligence for security-related activities as a benefit of collective sharing.

Sharing with a trusted analytics server while adhering to the company data sharing agreements could be particularly appealing and efficient. Since the assumption of a trusted server may not be valid, some solutions for privacy-preserving collaborative data analytics have been suggested in the literature, based on secure multiparty computation (SMPC), and more recently with homomorphic computing [7]. Through SMPC protocols, each player (party) contributes to the collaborative analysis process, conducting association rule mining without disclosing its private data to other parties [7]. This approach ensures a high-security level but strongly affects the system performance due to the computational requirement for the protocol that increases with the number of players and processed data, thus being not scalable. To mitigate the performance overhead, part of the workload can be given to a commodity semi-trusted server [7], which handles the operation independent from the private data (on sanitised/anonymised data set). This result accuracy degradation concept has been recently formalised in [8], where the first set of analysis operations is performed locally by the information providers. Afterwards, the results are sent to a central data mining entity for collaborative data analysis. The amount of information loss, which depends on the local operation, is formalised in a variable that affects the final result accuracy.

Since information sharing is a major issue our research work in the introduction of DSA and a platform to facilitate this information sharing.

## State of the Art

### Search Strategy and Inclusion Criteria

A systematic literature review was made by following PRISMA (Preferred Reporting Items for Systematic Reviews and Meta-Analysis) Methodology [9], and with the research question “What is the state of the art on how Health organizations share data for data analytics?”.

We have searcher databases such as Scopus and Web of Science Core Collection (WoSCC) and the research was conducted through March 26th, 2022; all the results had to be articles, published between 2017-2022 and written in English.

The search strategy was based on one query made with different focuses of research. This method allowed for the observation of the number of articles existing in both databases, considering the concept, context, population and domain under study.

This method allowed for the observation of the number of articles existing in both databases, considering the concept and context, and population under study. It is important to note that the values corresponding to the queries still have duplicate articles.

For this review only articles were considered. Grey literature, reviews, conference papers, workshops, books, and editorials were excluded, as well as works not related to the domain.

## Study Selection

Firstly, the selection of papers was done using the title and abstract, and in some cases in which that information was insufficient, the full document was analyzed.

### Data Extraction and Synthesis

The data were managed and stored by Zotero and Microsoft Excel. These data were title, author, year, journal, subject area, keywords and abstract. For data synthesis and analysis, a qualitative assessment was conducted based on the results presented above. All the databases – Scopus and WoS – were searched systematically regarding the published work related with the domain of “data analysis” or “data analytics”, the concept “Data Sharing Agreement*” or “Data Sharing”, the target population “Organization*” or “Patient*” or “Management Process*” and within a “e-health” or “health” context of the study.

## Results

The research was made by searching the existing literature regarding the concept, target population and the context of this study in Scopus and at the WoSCC detailed in Table 1. The query was made in the individual databases and with the same restrictions and filters (It is important to note that the values corresponding to the queries still have duplicate articles).

**Table 1.**
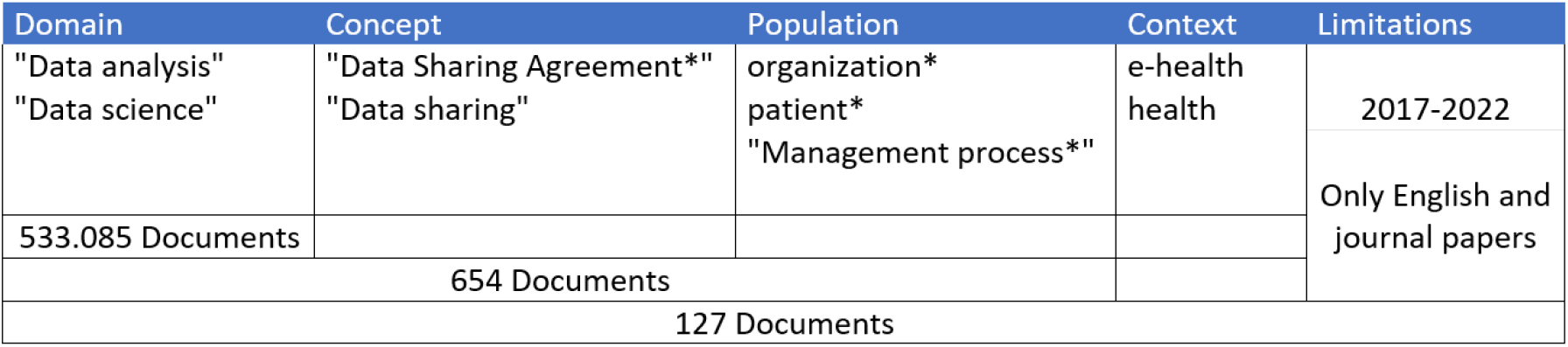
Research done on target population and the context of this study in Scopus and at the WoSCC

From this we can see that when the query is made using the keywords from each column (Domain AND Concept AND Population AND Context AND Limitations) returning 127 documents.

After performing a manual process, towards the identification of significant subjects on their research questions, identifying the outcomes and removing the duplicates, 41 documents were obtained. Our research systematization considered year, area, RQ topic and a small description.

Figure 1 shows the PRISMA workflow diagram from the total of articles studied.

**Figure 1.**
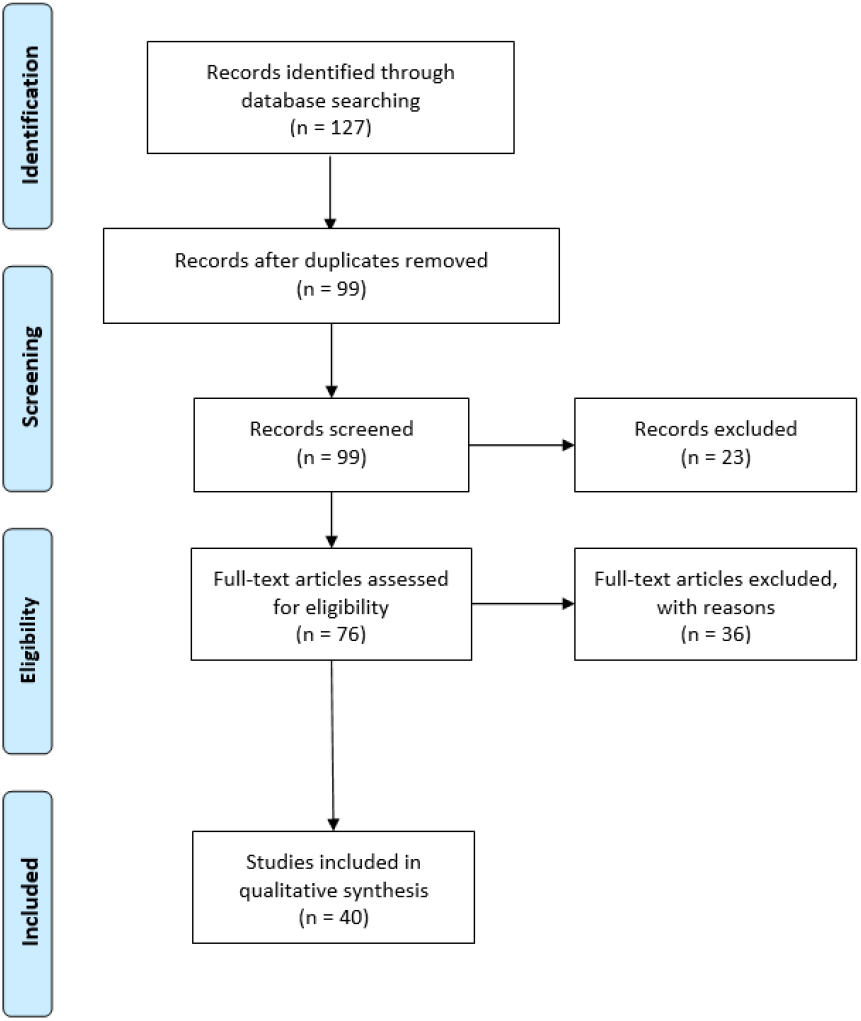
PRISMA methodology used in the literature revision

### Study Characteristics

All the 41 studies included in the review were selected through the use of the specific criteria mentioned above.

From Figure 2 we can notice from the trend line that there is a growth on the topic that we are studying, revealing his relevance. On this Figure, only completed years were included.

**Figure 2.**
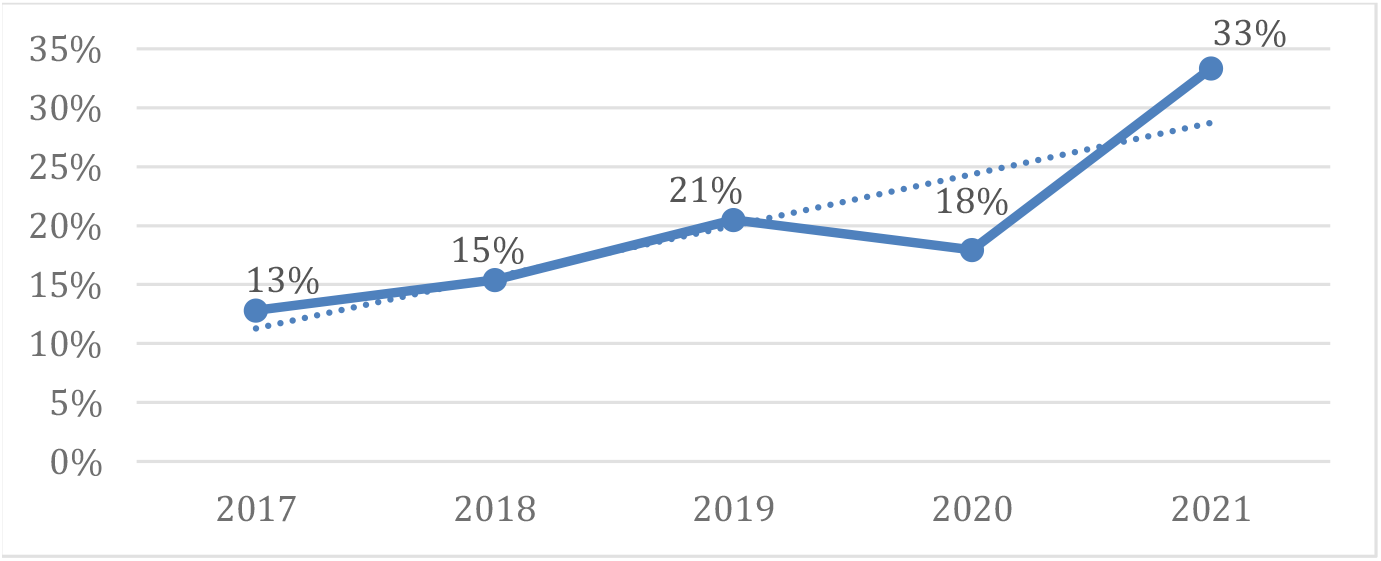
Percentage of publications in last 5 years

Considering the goals of this article is to identify the use of Data Sharing Agreements between Health Organizations, a list of the main topics discussed on each of the reviewed articles are described on Figure 3, where it is noticeable the focus on the Data Sharing concerning privacy and blockchain, being this the 3 main topics.

**Figure 3.**
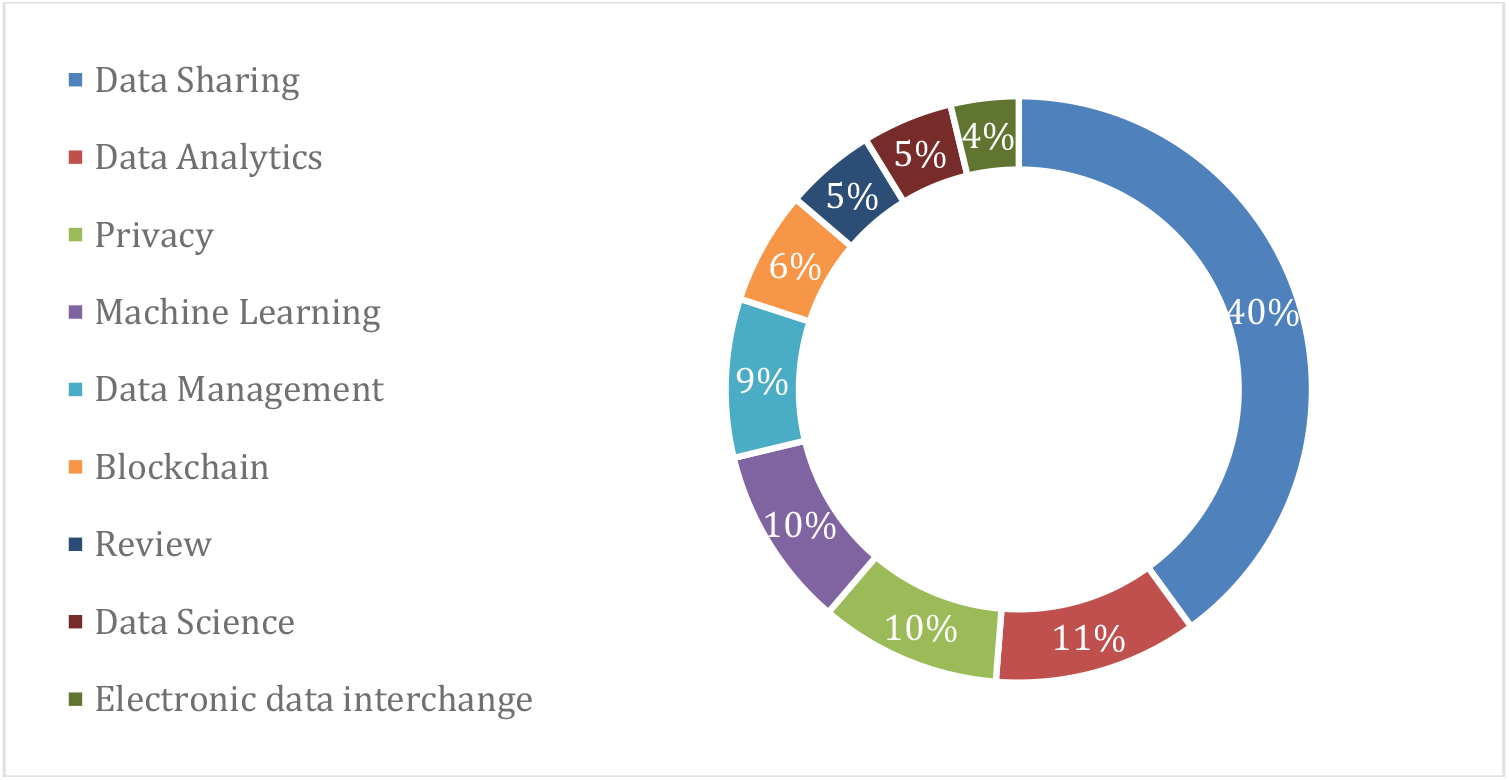
List of Topics Publicated

A more detailed analysis of this review is summarized in Table 2. The description of the topics was explicit and no requests for clarification were necessary to the authors of the articles.

**Table 2.** Detailed information about topics and related papers

| Topic | Reference |
| --- | --- |
| Review | [10]–[13] |
| Blockchain | [10], [11], [14]–[16] |
| Privacy | [10], [11], [13], [14], [17]–[20] |
| Electronic data interchange | [10], [13], [15] |
| Data Analytics | [21]–[29] |
| Machine Learning | [13], [20], [21], [25], [27], [30]–[32] |
| Data Management | [13], [18], [21], [33]–[35] |
| Data Science | [21], [30], [36], [37] |
| Data Sharing | [11], [12], [15]–[17], [19], [20], [22], [23], [27]–[33], [35]–[49] |

As mentioned before, the classification of the studies regarding the outcome is not mutually exclusive, given that these were attributed due to presence/absence in the study.

### Goals and Outcomes Analysis

The use of shared data is indeed the most common topic from the gathered studies. Concerning data sharing, the current study focusses on Data Sharing Agreements, so we will first do an analysis of the studies that imply data sharing. A common topic for this is blockchain, and study [11] presents a systematic literature review with the goal of analyzing the motivations, benefits, and limitations, as well as barriers and future challenges, when using distributed ledger technology in oncology, where the authors conclude that blockchain has the potential to improve data sharing (for primary care and medical research), as well as achieve pharmaceutical supply chain optimization by bringing properties such as transparency, traceability, and immutability to the table. Authors on study [14] consider how blockchain technology might help with data exchange via different types of mechanisms, concluding that It’s still unclear if blockchain can help with the shift from institution-centric to patient-centric data exchange. Glicksberg et. Al on study [15] created and tested a blockchain-authenticated system for secure sharing of deidentified patient data derived from standard of care imaging, genomic testing, and electronic health records (EHRs), demonstrating the system’s viability as a framework for sharing health data trapped in silos to advance cancer research. Study [16] demonstrates the potential of blockchain to stimulate successful healthcare data sharing while ensuring the security of original data sources with four contributions to the research of using blockchain technology to clinical data sharing in the context of technical needs. Moving away from blockchain, study [43] introduces a Data Warehouse, as the first multicenter for electronic health records with full admission data from COVID-19 patients, using data sharing agreements between hospitals showing that these activities are critical for the advancement of medical data science in acute care medicine.

Concerning privacy issues, study [10] reviews the state-of-the-art schemes for safe and privacy-preserving medical data sharing throughout the last decade, with an emphasis on blockchain-based techniques, concluding that there are still certain issues that require more research and study in blockchain-based medical data management. Authors from [13] review and discuss required technique contributions to help multisite EHR data networks share data more easily. On [17] suggest a fog-assisted health data exchange mechanism for e-healthcare systems that is both efficient and private. To preserve privacy, study [19], by using attribute-based encryption and identity-based broadcast encryption approaches, achieve secure and fine-grained health data and social data sharing, allowing patients to securely exchange their sensitive personal data, whereas study [20] To keep data useful while maintaining privacy, employed a risk-based deidentification technique. Blending the privacy and data management topic, study [18] provides a protocol for a project aimed at coproducing a people-centered paradigm for including patients and the public in decision-making processes around the use and sharing of health data for rare illness treatment and research.

For topics such as data analytics using data sharing, authors on [22] argue that employing digital health technology to facilitate the pooling of patient data from diverse sources for research and regulatory reasons has a lot of promise. Study [23] data sharing facilitates acute medical research by establishing a foundation for new studies and disseminating data, pictures, and biomaterials for future study. On [27] established an architecture that demonstrates that patient privacy hurdles to healthcare data sharing can be overcome and that quick data analysis can be performed across several institutes from different countries with varied legislative regimes. Zhang et al. on [28] offer a data library of consistently processed genomic and related clinical data to the cancer research community, allowing data sharing and collaborative analysis in support of precision medicine. Authors on [29] have the goal to introduce and discuss Medical-Blocks, a platform for exploring, managing, analyzing, and sharing data in biomedical research via a file hosting service for collaboration, as such as been demonstrated to be needed to enrich studies that lack medical imaging data [50]. On image, study [30] identifies five major fields of activity crucial to cooperation with patient data: privacy, informed permission, standardization of data pieces, vendor contracts, and data valuation, and proposes philosophies around best practices in the sharing of health information.

On the data analytics theme, authors from [21] seek to make health care and medication research more efficient and focused by utilizing machine learning to handle enormous amounts of anonymized data from a variety of data sources and kinds, with the goal of identifying unique patterns with clinical value that cannot be recognized by humans alone.

Data sharing is a crucial part of improving the health-care system [34] and it is very important in a post-pandemic scenario, as shown by study [33], where this data can be used to assist families in locating the graves of family members who died during the pandemic and are the finest tactics in outbreak response.

Data is necessary for providing ethical, lawful, safe, and efficient direct care, service planning and improvement, and research [35]. On this study have created a system that enables for safe, regulated electronic exchange of a person’s health information between systems and healthcare organizations in order to improve healthcare quality and efficiency.

## Methods

We employed an action research strategy, which is appropriate when the goal is to effect change in real-world settings [51],[52, pp. 14–32]. Action research aims to solve a particular issue in a specific situation. Contributing to both practice and analysis requires the combined experience of practitioners and researchers, which necessitates a collaborative research strategy [19].

The proposal baseline is that the Information Prosumers want to cooperate by setting up a dynamic federation to provide their information to (a set of) data analytic services that will be able to detect security threats that would not be discovered exploiting the data of one prosumer only.

The proposed platform for providing such collaborative information consists of two main elements: the Information Sharing Infrastructure (ISI), which allows the sharing of information among the member of the federation while protecting information confidentiality, and the Information Analytics Infrastructure (IAI), which implements the specific data analytics services (the results are then stored in the ISI for further processing). This can be combined in a P2P (pear to pear) structure allowing several layers of refined analysis (e.g., in a tree-like facility).

In this context, protecting the information provided by the parties belonging to the federation is a primary issue. The proposed platform will allow Information Prosumers to set up digital agreements concerning the usage of their information and the results produced from them. The platform will enforce these agreements before and after the execution of the analytic functions.

Hence, the proposed approach (see Figure 1) allows the Information Prosumers to 1) share their information only with a given subset of members of the Federation; 2) decide which of the available analytics operations can be executed on their information; 3) perform some pre or post-processing manipulation operation, which must be executed on their information; and 4) decide to disclose the analysis results only to certain Information Prosumers and under certain conditions.

The digital agreements among prosumers concerning the usage of their information, called Data Sharing Agreements (DSAs), are defined when the prosumers create the Federation and were established based prosumers interests. A DSA is an agreement between two or more parties who wish to exchange data in several specific domains and contexts: it regulates which data to use, for which purposes and how to use them. DSA aims to capture the data sharing policies that restrict both suppliers and consumers of data and govern the flow of data between them. The ISI grants continuous enforcement of the policies and its obligations. In this scenario, DSAs are concerned both the prosumers’ information and the data derived from the analytics computation. The DSA also regulates the storage of information. In particular, DSAs express constraints on the shared information in terms of 1) manipulation operations (obligations on): to pre-process the information before or after its usage, e.g. to anonymise the data before processing or before sharing the analysis results with the federation (or even with a specific member); in particular, we consider anonymisation and homomorphic computing operations; and 2) analytics operations (authorisations on): to permit or not an analytic service depending on certain conditions, e.g. “Detect Inactive User Activity” service is authorised only after the execution of specific manipulation operation on the data like “Data must be anonymised.”

Hence, the DSAs allow prosumers to define which manipulation operations must be performed on the shared information before the elaboration of the analytics engine, which analytics operations can be performed on the manipulated data, and which manipulation operations must be performed on the results before sharing them with the members of the Federation. It is worth noticing the distinction between manipulation and analytics operations since manipulations operations concern several information preparation operations (defined in specific enforceable policies) executed on the information of one single prosumer mainly for preserving confidentiality and privacy, such as data (pseudo) anonymisation, etc. Conversely, analytics operations consist of all the computation operations performed on the information shared by all the prosumers to extract the relevant information.

The information workflow in the proposed architecture is the following: 1) An Information Prosumer sends his information to the Information Analytics Service (IAI) through the Information Sharing Infrastructure (ISI); 2) The ISI executes the manipulation operation specified in the DSA related to this information before the information is sent; 3) The manipulated information is sent to the IAI along with the related DSA; 4) The IAI enforces again the DSA paired with the information by performing further manipulation operations, checking that the requested analytics operation is allowed by the DSA, and executing a manipulation operation on the results before sharing with the federation members; and 5) The results are returned to all federation members, who can take their actions as a consequence.

The Information Sharing Infrastructure (ISI) (see Figure 2) is a virtual layer that is deployed when a set of Information Prosumers set up a federation by defining Data Sharing Agreements (DSAs) to share their information. The Information Sharing Infrastructure is in charge of managing the Federation of Information Prosumers by allowing them to define their DSA, collecting data from these prosumers, by enforcing the DSA paired with the information before the execution of the analytics operations, by retrieving the results computed by the Information Analytics services and distributing them back to the Information Prosumers again enforcing the DSAs to respect the confidentiality and privacy requirements of each of them. The ISI’s main components are the DSA enforcement engine and the data protected object store (data are encrypted in rest and with appropriate usage policies). The Information Analytics Infrastructure (IAI) (see Figure 3) is in charge of providing specific data analytics services, also deployed as plugins, and allowing the development of efficient and reliable execution of these services. The overall IAI will allow: 1) Efficient deployment on Data Analytics Operations (possibly as plug-ins) enabling Information Analytics Service trusted market; 2) Enforcement, in cooperation with the ISI, of the DSAs; 3) Providing specific data analytics operations such as clustering, classification and data correlation for our Pilots; and 4) Providing efficient, privacy-aware and reliable distributed computation service.

The IAI receives data from ISI. Data are analysed through the collaborative analysis service, which includes tools for parallel computing, sharing the computation between several machines to allow fast computation on a very large amount of information. The IAI offers toolboxes for analytics based on Machine Learning which can be exploited for intrusion detection and attack pattern recognition, Deep Learning which can be used for seamless authentication, image and sketch analysis and Statistical correlation, which includes, among the others, prediction services, cascade analysis to correlate vulnerabilities to threats and attacks which might exploit them. The collaborative analysis service uses machine learning suites, including classification, clustering and statistic algorithms to extract additional knowledge relevant to the prosumers. The new computed information is finally returned and eventually distributed to the Prosumers under the DSA stated conditions.

The IAI also exploits privacy-aware analytics techniques, particularly homomorphic encryption, which complete the set of security operations offered by the DSA manager. These privacy-aware analytics techniques allow collaborative analysis on encrypted data, allowing the enforcement of security policies where the analytics platform is not considered trusted.

Through this approach, prosumers (e.g., Hospitals) can invoke and use a trusted and interoperable set of data analytics services accessible through a market-like platform. This demands standardisation of core elements of the approach framework (e.g., Data Analytics Operations) and definition of clear trust boundaries based on security certification schemes. It is worth noticing that among the benefits of setting up data prosumers collaboration, we can list: 1) benefits coming from a single Prosumer are promptly shared, under DSA conditions, between all other prosumers registered to the service; and 2) possibility to correlate events, mainly related to security, co-occurring in the infrastructures of different prosumers that would otherwise go unnoticed.

The approach elicitates the specific requirements for each stakeholder and ensures that the project will design and implement dashboards that are tailored to different roles and users, being able to derive multiple sets of dashboards that are aligned with the identified information needs, see Figure 4. In this sense, the different dashboards to be implemented will need to be adapted according to (i) the tasks and responsibility assigned to each actor (i.e. the relevant information they should be aware of), (ii) their particular perspective of the ecosystem (i.e. what are they interested in), and (iii) which other actors they interact with (i.e. relevant patients, caregivers, etc.). To this aim, the implementation of the dashboards will be carried out by designing the initial mock-ups for the dashboard set and then iteratively interviewing the interested parties as dashboards are implemented and refined. This dashboard allows advancing proficiency in data-oriented health services. Analysing Big Data requires extensive use of visualisations. Visualisations play a crucial role in discovering and transforming data into knowledge, especially when dealing with large volumes such as those found in Big Data scenarios].

**Figure 4.**
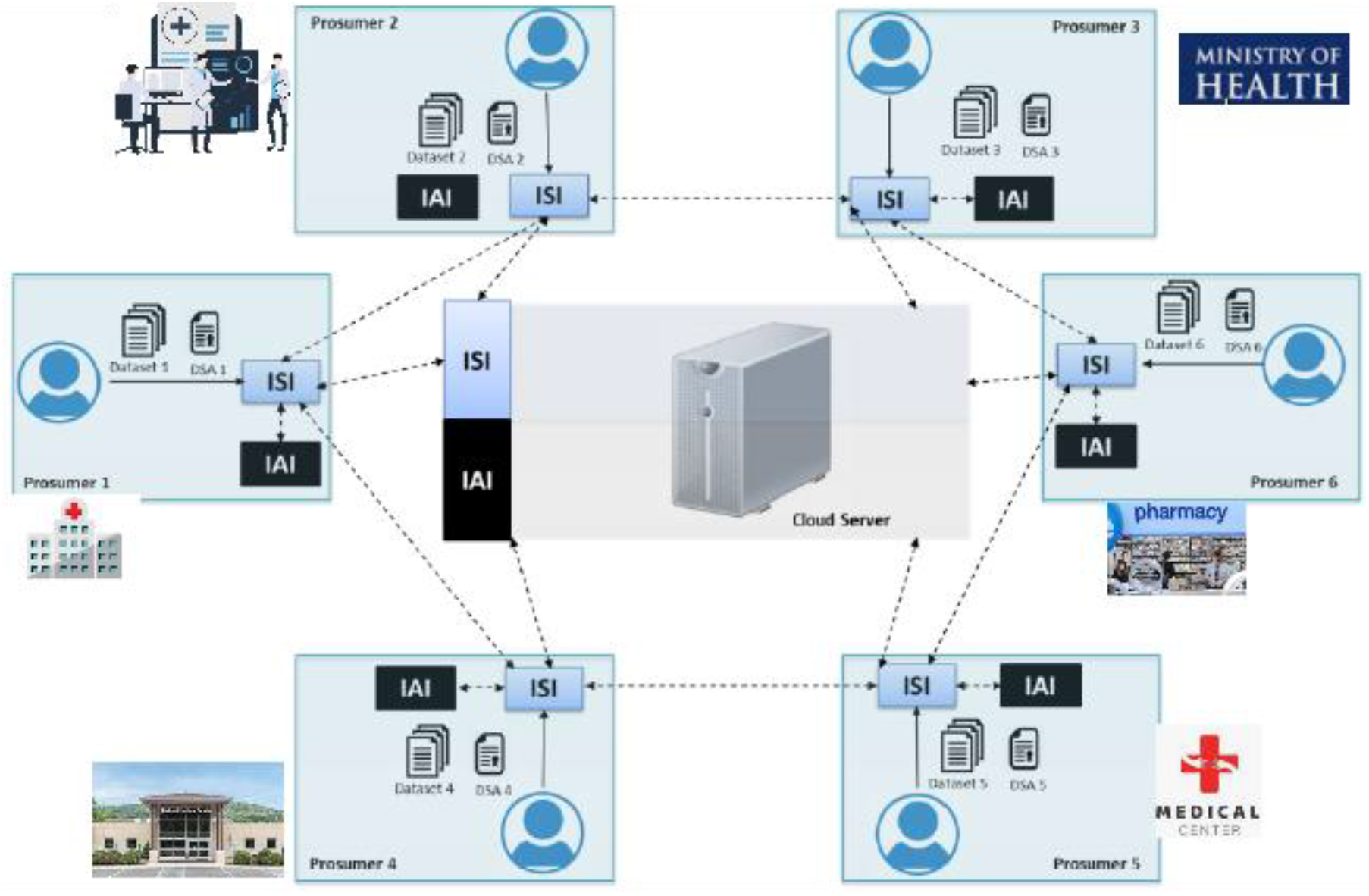
Framework architecture

**Figure 5.**
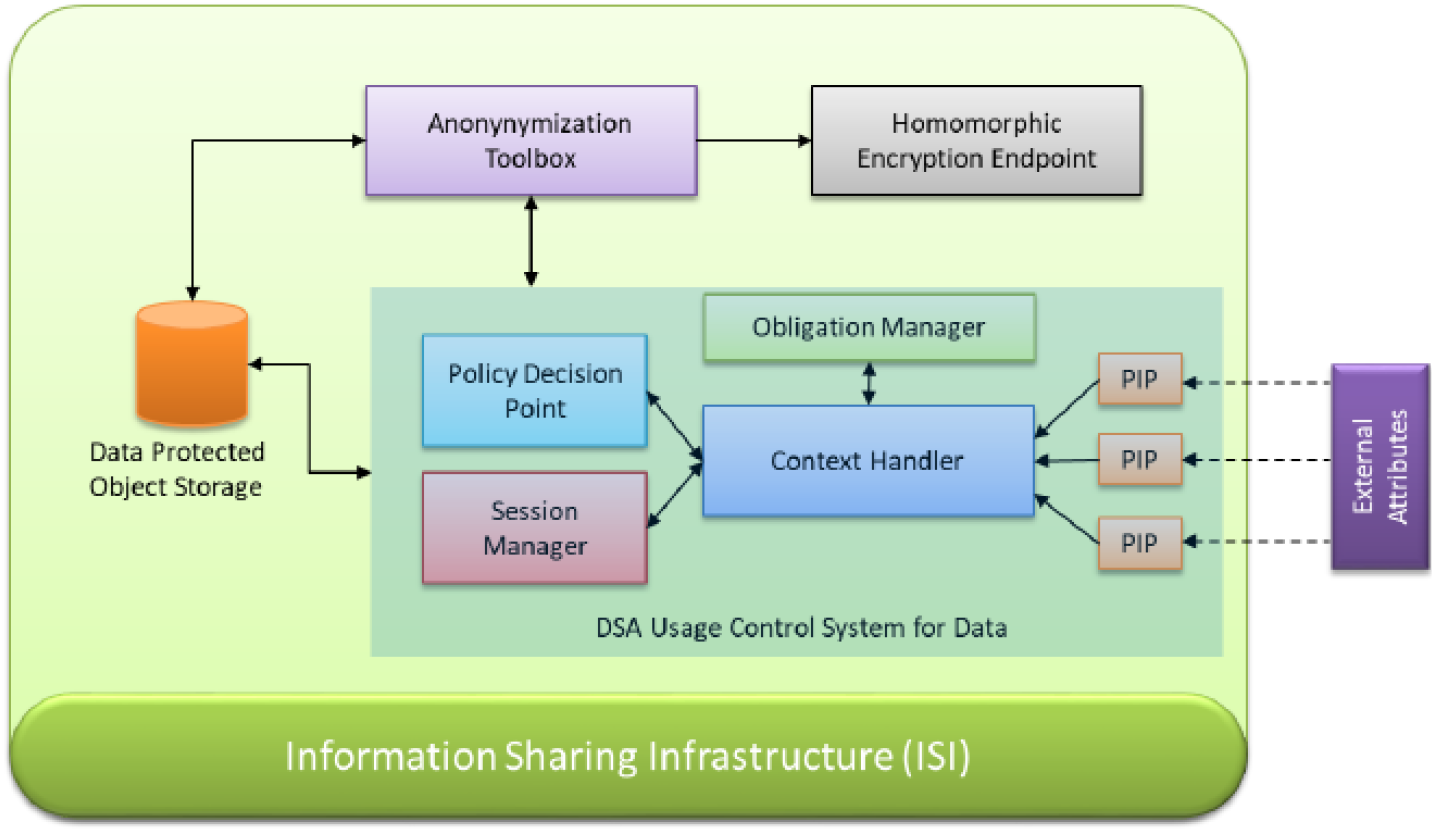
Information Sharing Infrastructure (ISI)

**Figure 6.**
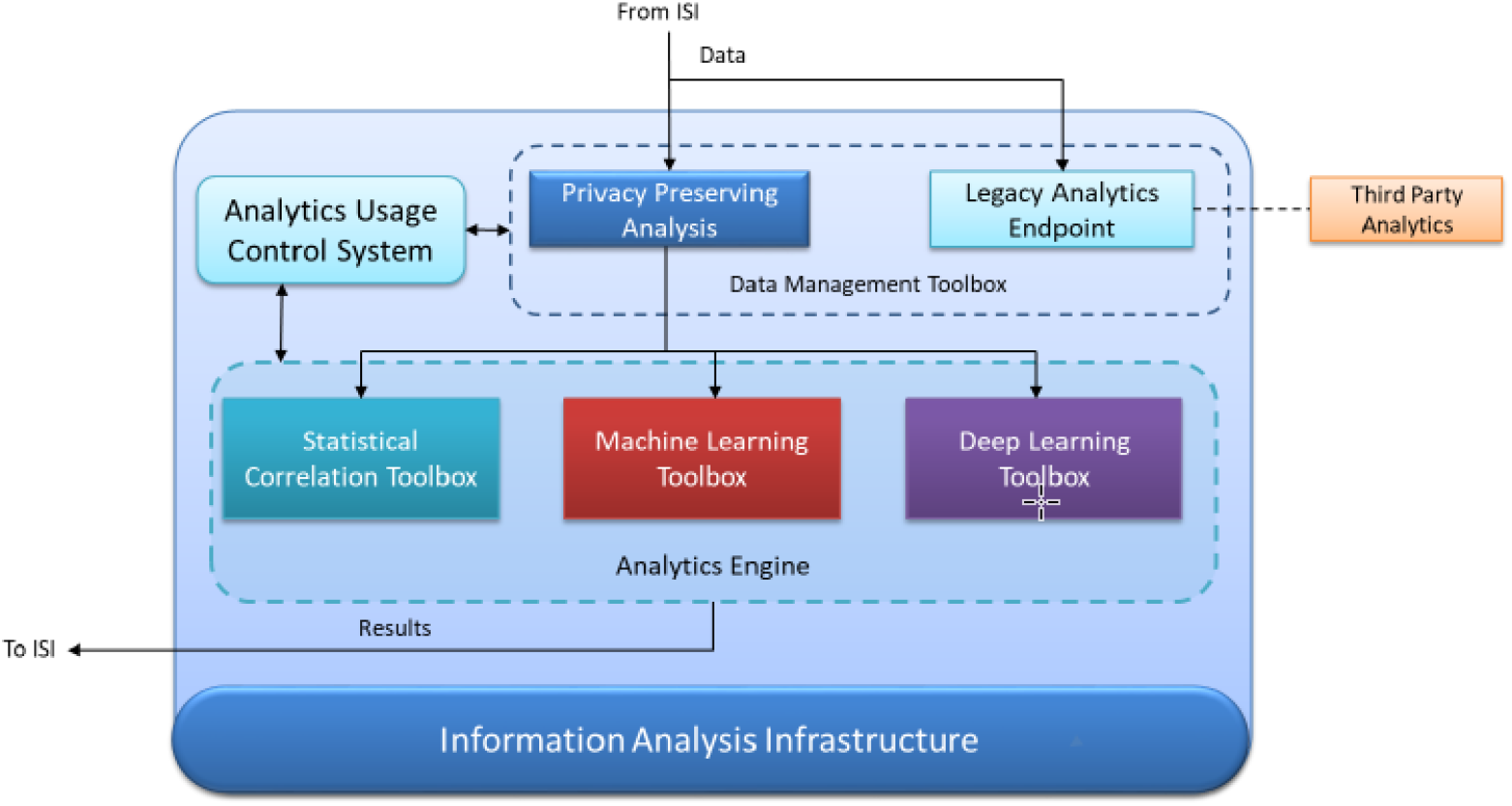
Information Analysis Infrastructure (IAI)

**Figure 7.**
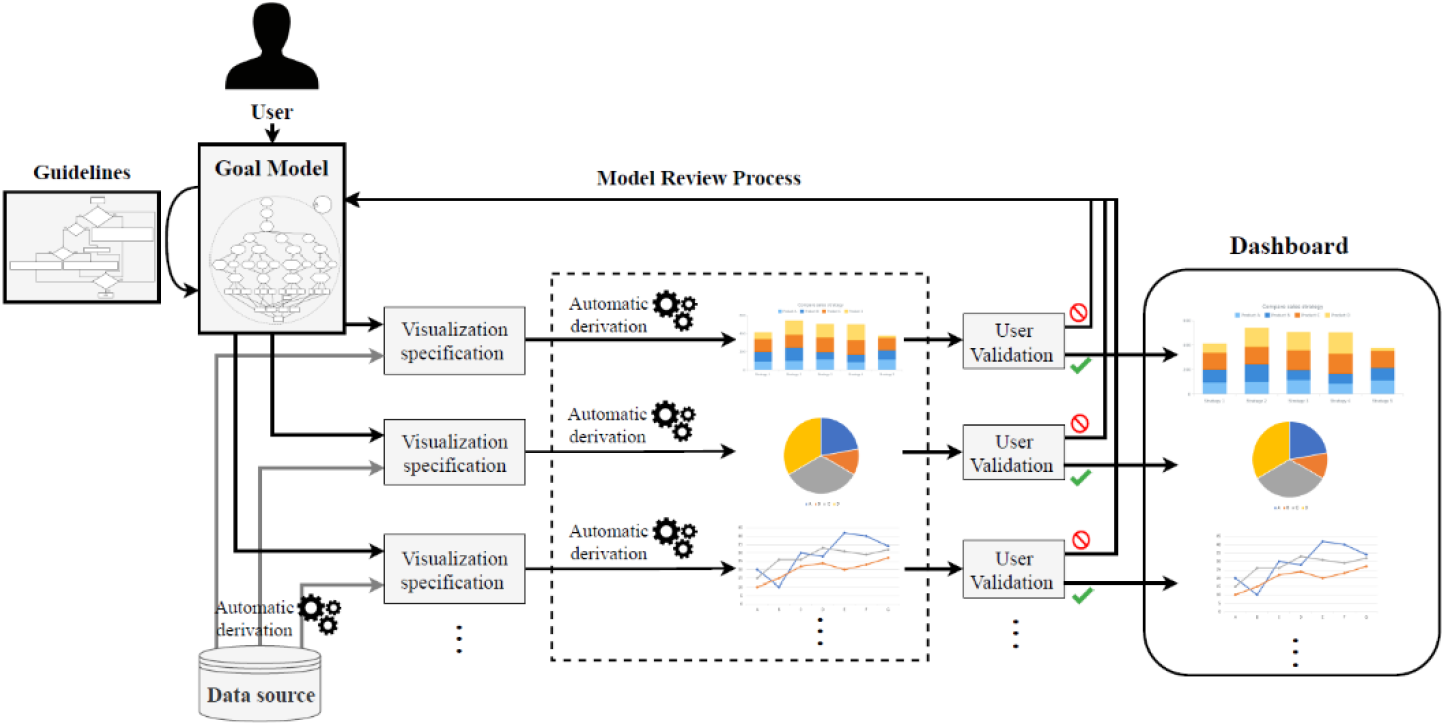
Dashboard conceptualisation, design and implementation methodology

## Conclusions

The framework enables the rapid and successful implementation of contractual agreements between businesses/entities that offer and consume data. This is done in a secret and privacy-preserving manner, consistent with both the company’s data policies and the requirements of current data privacy law (i.e., the European data protection directive 95/46/EC and any future amendments). The system imposes constant data use management and data security through encryption (in rest and transit).

The platform as a whole provides data analytics as a service in a secure and collaborative manner. Numerous technologies will be used to construct this data analytics service, depending on the amount of confidence that prosumers place in the services, i.e. whether the services are trusted or not. The most suitable data protection and analysis procedures were created in order to trade-off several aspects, such as privacy vs analysis accuracy. The framework will be efficient and safe, and it will be an open platform with an open API that will facilitate the framework’s integration and adoption.

The platform specifically analyzes data analytics tools for security (including log and behavioral analysis), the use of homomorphic computing for chosen data analytics tasks, data anonymization methods for data sanitization, data analytics visualization tools, and managed security services.

To author’s knowledge this is one of the first approaches in DSA to share information in the health sector. Other approach to share information is through the blockchain but in this case the shared AI services are more complex to implemented. This research aims at providing a flexible, secure and privacy-aware framework allowing confidential, distributed information sharing in health entities. Information sharing in the health sector is proposed based on the digital service agreement, interface to the local system and holomorphic encryption (HE) to allow sharing of Artificial Intelligence (AI) services among different health stakeholders. This allows knowledge creation based on shared services and digital services agreements (DSA) is one first step towards data and information sharing in the health sector. We implemented an information analysis infrastructure using DSA, as a concept proof, with health system connectors and a set of AI services using HE.

Major limitations are regarding the adoption of DSA and populate the system with information

## Data Availability

no

## Acknowledgements

This work is partially funded by national funds through FCT—Fundação para a Ciência e Tecnologia, I.P., under the project FCT UIDB/04466/2020 and UIDP/04466/2020. Luís Elvas holds a Ph.D. grant, funded by FCT with UI/BD/151494/2021

